# Using Genome Sequence Data to Predict SARS-CoV-2 Detection Cycle Threshold Values

**DOI:** 10.1101/2022.11.14.22282297

**Authors:** Lea Duesterwald, Marcus Nguyen, Paul Christensen, S. Wesley Long, Randall J. Olsen, James M. Musser, James J. Davis

## Abstract

The continuing emergence of SARS-CoV-2 variants of concern (VOCs) presents a serious public health threat, exacerbating the effects of the COVID19 pandemic. Although millions of genomes have been deposited in public archives since the start of the pandemic, predicting SARS-CoV-2 clinical characteristics from the genome sequence remains challenging. In this study, we used a collection of over 29,000 high quality SARS-CoV-2 genomes to build machine learning models for predicting clinical detection cycle threshold (Ct) values, which correspond with viral load. After evaluating several machine learning methods and parameters, our best model was a random forest regressor that used 10-mer oligonucleotides as features and achieved an R^2^ score of 0.521 ± 0.010 (95% confidence interval over 5 folds) and an RMSE of 5.7 ± 0.034, demonstrating the ability of the models to detect the presence of a signal in the genomic data. In an attempt to predict Ct values for newly emerging variants, we predicted Ct values for Omicron variants using models trained on previous variants. We found that approximately 5% of the data in the model needed to be from the new variant in order to learn its Ct values. Finally, to understand how the model is working, we evaluated the top features and found that the model is using a multitude of k-mers from across the genome to make the predictions. However, when we looked at the top k-mers that occurred most frequently across the set of genomes, we observed a clustering of k-mers that span spike protein regions corresponding with key variations that are hallmarks of the VOCs including G339, K417, L452, N501, and P681, indicating that these sites are informative in the model and may impact the Ct values that are observed in clinical samples.

## Introduction

The COVID-19 pandemic, caused by severe acute respiratory syndrome coronavirus 2 (SARS-CoV-2), has had an extreme public health impact over the last three years. Since its emergence, it has caused over 625 million confirmed cases and over 6 million deaths worldwide (1). The SARS-CoV-2 virus has evolved over time to become more transmissible, resulting in new variants of concern (VOCs) causing successive waves of infection (2, 3). This sequential and ongoing emergence of VOCs, such as those observed in late 2020 with Alpha, followed by Delta, and continuing into 2022 with Omicron (4), presents a substantial public health threat. Despite this, the identification of new VOCs usually occurs only after there is community transmission of the new variant, hampering efforts to control viral spread (5, 6).

Clinical testing has played an important role in the pandemic response, enabling early identification and intervention. Real-time reverse-transcription polymerase chain reaction (RT-PCR) tests are the gold standard molecular diagnostic for detecting SARS-CoV-2 (7). The viral RNA in a patient sample, most commonly collected via a nasal swab, is converted to DNA by reverse transcription, and then amplified via PCR until the resulting SARS-CoV-2 cDNA is detectable. The Cycle Threshold (Ct) value of a positive SARS-CoV-2 test is the number of rounds of PCR amplification that are necessary for the amplified sequence to reach the point where it becomes detectable by the clinical detection instrument. Although there are many factors that can influence the Ct value observed in a clinical test including time since infection, sample preparation and quality, and differences in detection reactions and instruments (8), Ct values tend to be inversely correlated with viral load, providing a useful approximation of the viral RNA in a patient sample (9–11).

Ct values can also serve as a valuable source of epidemiological data. For example, Ct values for cross-sectional samples collected from patient populations over a given time period are often indicative of the state of the epidemic, with lower average Ct values indicating a growing pandemic (12–14). Similar studies have also found that cross-sectional trends in Ct values can act as indicators of the future trajectory of the pandemic (15, 16). At the patient level, Ct values have been shown to provide a good estimate of how long a patient will remain contagious (10, 17, 18), and several studies have shown that lower Ct values (i.e., higher viral loads) can be correlated with symptomatic infection, morbidity, and mortality (9, 14, 19, 20). Higher viral loads and increased transmissibility have been reported for each successive VOC as it has emerged, including the Alpha, Delta, and Omicron variants (21–23). Several mutations found in these VOCs, such as D614G and P681R in the spike protein, are linked with increased viral loads and lower Ct values (24–26).

Because VOCs significantly alter the course of the pandemic and threaten the efficacy of current treatment methods, developing tools for the early identification of mutations that could lead to increased transmissibility is important. Such early identification could enable prompt clinical responses to greatly limit, or even prevent, the spread and effect of the new variant. Genomic surveillance studies have aggregated data from public repositories to develop models to identify mutations involved in transmissibility and identify notable variants with the potential to spread (6, 24, 27, 28). Similarly, studies analyzing whole genome and individual protein sequences using statistical and machine learning methods have also been developed to predict disease severity (29), vaccine targets (28, 30), transmissibility (27), and future variants of concern (31–33).

Although the published literature is indicating that Ct values for positive SARS-CoV-2 samples are valuable epidemiological metadata, they are rarely reported with deposited genome sequences. This has subsequently limited their use as features in predictive modelling studies. Over the course of the pandemic, the Houston Methodist Hospital System has been attempting to sequence every positive SARS-CoV-2 clinical sample from their centralized diagnostic laboratory (34–38). In this study, we use over 29,000 high quality SARS-CoV-2 genome sequences from this collection, paired with their Ct values from clinical detection instruments, to determine if machine learning models built from the genome sequences can predict Ct values and indicate important mutational sites influencing viral loads.

## Materials and Methods

### Data collection

A total of 29,272 SARS-CoV-2 genome sequences were used in this study. The sequences were collected across the Houston Methodist Hospital system and from institutions using the Houston Methodists diagnostic laboratory services between May 15, 2020 and January 14, 2022. Nearly all samples were collected from nasopharyngeal swabs immersed in universal transport media. Oropharyngeal or nasal swabs, bronchoalveolar lavage fluid, and sputum treated with dithiothreitol were used in rare cases. All samples were collected with patient consent and Institutional Review Board approval from the Houston Methodist Research Institute (IRB1010-0199). Positive clinical samples and Ct values were generated on multiple clinical detection platforms. The dataset in this study is comprised of Ct values from three detection systems: the Abbott Alinity *m* SARS-CoV-2 AMP kit (Abbot Molecular Inc., Des Planes, IL, USA), the SARS-CoV-2 Assay using the Hologic Panther instrument (Hologic, Marlborough, MA), and the Xpert Xpress SARS-CoV-2 test using Cepheid GeneXpert Infinity or Cepheid GeneXpert Xpress IV instruments (Cepheid, Sunnyvale, CA).

Upon clinical detection, samples were amplified for sequencing using either the ARTIC V3, V4, or V4.1 primers (V4 for collection dates after July 28, 2021, and V4.1 for collection dates after Jan 5, 2022)(https://artic.network/ncov-2019) using methods described previously (34–36) (**Table S1**). Nearly all of the genomes were sequenced using an Illumina NovaSeq 6000 instrument. Two hundred sixty-eight samples were sequenced on an Illumina HiSeq, and 989 of the samples were sequenced using an Oxford Nanopore GridION instrument. Genomes were assembled with the BV-BRC SARS-CoV-2 assembly service (39)(https://www.bv-brc.org/app/ComprehensiveSARS2Analysis), which performs a reference-based assembly against the Wuhan-Hu-1 reference genome (GenBank ID: MN908947.3). The pipeline uses minimap version 2.143 (40) for aligning reads against the reference and iVar version 1.2.2 for primer trimming and SNP calling (41). Default parameters were used in all cases except that the maximum read depth in mpileup (42) was limited to 8,000, and the minimum read depth was set to 3 in iVar. All variants were identified using Pangolin version 3.1.17 (https://cov-lineages.org/resources/pangolin.html) with pangolearn (2022-02-02).

### Genome quality filtering

The set of 29,272 genomes, each sampled from a unique patient, was down selected from a larger set of approximately 70,000 sequenced genomes in order reduce the effects of ambiguous nucleotide calls on the models. To do this, the average read depth at every position in the Wuhan-Hu-1 reference genome was computed across the larger set of genomes, and any position with an average depth lower than 10 was masked with an N character. The depth 10 cutoff was chosen by evaluating models with depth masking ranging from 0 to 40 and choosing the cutoff that maximized masking and minimized the loss in model accuracy (**Table S2**). This resulted in the masking of 56 positions in in every genome. These included positions 22029-22033, 22340-22367, 22897, 22899-22905, and 23108-23122, which correspond with spike amino acid positions 156, 157, 260-269, 445-448, and 516-520. The low average coverage in this region of spike is likely the result of poor priming. The first and last 100 nucleotides in each genome were also masked to prevent jagged edges from creating variation that could be incorrectly learned by the models. Overall, this resulted in a total of 256 masked positions in each genome sequence. If any genome had greater than 500 ambiguous characters in addition to this initial set of 256 masked positions, it was discarded.

### Feature matrix construction

In order to train a model on SARS-CoV-2 genome sequences, a matrix of nucleotide k-mers was constructed consistent with the methodology of Nguyen et al. (43). Briefly, the frequency of unique overlapping nucleotide 10-mers was computed for every genome using the k-mer counting program KMC (version 3.0.0) (44). KMC considers each strand of the k-mer and retains the lexicographically highest “canonical” k-mer. A matrix was constructed with features consisting of the frequency of all unique 10-mers occurring across all genomes. Each row of the matrix represents one viral sample and contains the frequencies of each 10-mer (0 if the 10-mer is not present in the genome) and a one-hot encoding representing the detection instrument.

### Model generation and evaluation

Unless otherwise stated, all models were generated using random forest regression from scikit-learn (version 1.0.2) (45) to predict Ct-values from the k-mer matrix. Random forest regression was chosen based on a comparison with XGBoost and logistic regression using default parameters and a set of informed parameters for each method (**Table S3**). A 2k factorial design was used to determine the importance of four hyperparameters in the random forest models: number of trees, tree depth, row subsampling, and column subsampling (46) (**Table S4**). Three hyperparameters were found to be important: number of trees, tree depth, and row subsampling. Hyperparameter tuning was carried out via grid search to identify the optimal values for each hyperparameter (**Table S5**). The final hyperparameters chosen for the random forest model were: none for tree depth, 400 for number of trees, and 0.25 for row subsampling, with all other parameters set at the defaults. Weighting the model by frequency of Ct values or variants was not chosen because it did not result in statistically significant differences in model performance (**Table S6**).

Following parameter tuning, a random forest model was built from all genomes using all variants and instruments. This model was evaluated through five-fold cross validation with a train-test-split of 80% and 20%. For each fold, the R^2^ score and root mean square error (RMSE) were computed.

Additionally, the model accuracy within Ct-value intervals of 6, 5, 4, 3, 2, and 1 were calculated. The accuracy within a given interval *n* was computed as the fraction of the samples where the absolute difference between the predicted and actual value was smaller than _n._ Unless otherwise stated, results are shown as the average across all five folds with 95% confidence intervals.

### Predicting Ct values in new variants

To assess the model’s ability to predict the Ct values for a newly emerging variant, we varied the amount of Omicron sequences in the training set to see how well models built from previous variants could predict Omicron Ct values. For each fold, a separate model was trained using training sets with the following percentages of Omicron genomes: 0%, 0.5%, 1%, 2%, 5%, 10%, and 15%, and 5-fold cross validations were computed. For each percentage, *n*, the corresponding number of Omicron genomes were removed from the training set to produce a training set consisting of *n*% Omicron genomes. The test set remained unchanged for all percentages across each fold.

The model trained on a training set of *n*% Omicron genomes was evaluated on 3 separate subsets of the test set, 1) all Omicron genomes in the test set, 2) all non-Omicron genomes in the test set, and 3) a test set consisting of *n* percent Omicron genomes (generated using the same methodology as the training set). These experiments evaluated different aspects of the performance of a model trained on increasing amounts of a new variant: 1) how well a model could learn to predict a new variant, 2) whether including a new variant in the training set could interfere with the model’s ability to predict other variants, and 3) the model’s overall accuracy.

### Feature importance

The “feature_importance” function for random forest was used to analyze important k-mers for the all-instrument model. This returns the Gini impurity at each decision node (k-mer). Since rarely occurring features can have low impurity, we also analyzed important features by computing the decision paths from each tree in the forest for each genome (one decision path for each of the 400 trees in the forest, per genome). In this way, we defined an important feature as a node that is encountered most frequently for all trees and genomes. The most frequently occurring k-mers were analyzed in two ways. First, all k-mers where there was a significant difference in the average Ct values of the genomes with and without the k-mer were retained. Significantly different sets were defined as those with non-overlapping 95% confidence intervals for the average Ct values with and without the k-mer. Then for all k-mers in this set, a baseline was established empirically in order to display those k-mers that occurred most frequently relative to the background. In the second approach, the same set was also computed based on the criteria above, without the baseline filtering, and instead showing the set of k-mers that occurs in at least 10% of the genomes. The coordinates of the top k-mers were calculated based on their nucleotide alignments against the Wuhan-Hu-1 reference genome (GenBank ID: MN908947.3) using Mafft v7.475 (47) with default parameters.

## Availability

The code for the Ct value prediction model and the documentation for running the code and model are available at the following GitHub page: https://github.com/BV-BRC-dependencies/Ct-Value-Prediction. All genome accessions are provided in **Table S1**.

## Results

### Models built from sequence data are predictive

A total of 29,272 SARS-CoV-2 clinical samples, collected over a period of 19 months from May 15, 2020 to January 14, 2022, were used in this study. All positive samples were identified on one of three clinical detection instruments including Alinity (22,271 samples), Panther (6,418 samples), and Cepheid (583 samples) (**Figure 1**). The corresponding high quality sequenced genomes contain 324 different variants of SARS-CoV-2 with 106 variants occurring 10 or more times. The most common variants during the sampling period were Delta variant and its sub-lineages (6,285 samples), Omicron and its sub-lineages (5,445 samples), the related B.1(3,161) and B.1.2 (4,839 samples) variants from early in the pandemic, and Alpha (B.1.1.7) (1,478 samples). The Ct values in the dataset ranged from 5.1 to 44.9 with a median of 24.9, and a standard deviation of 8.2 (**Table S1**). The distributions of Ct values over the set of samples are similar for Alinity and Panther; the Cepheid distribution differs slightly, although it has a small number of samples (**Figure 2**). The Cepheid samples also had slightly higher median Ct values (28.5) versus the samples from the instruments (24.8 and 24.7 for Alinity and Panther, respectively) (**Table S7**).

**Figure 1.**
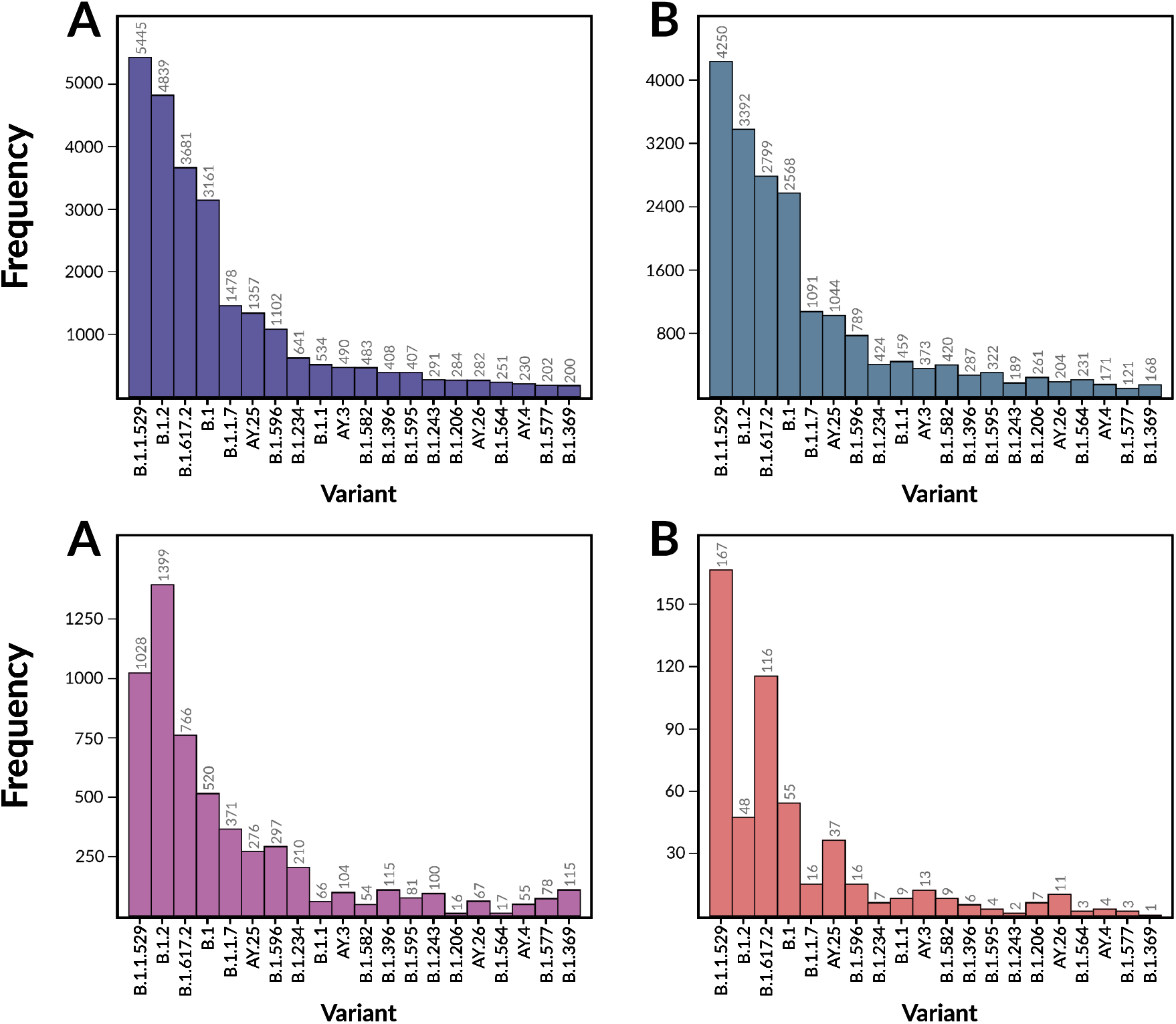
Histograms showing the distributions of the 20 most common variants in the dataset. **A**: The distribution of variants across all detection instruments. **B**: The distribution of variants in the genomes from the Alinity detection instrument. **C**: The distribution of variants in the genomes from the Panther detection instrument. **D**: The distribution of variants in the genomes from the Cepheid detection instruments.

**Figure 2.**
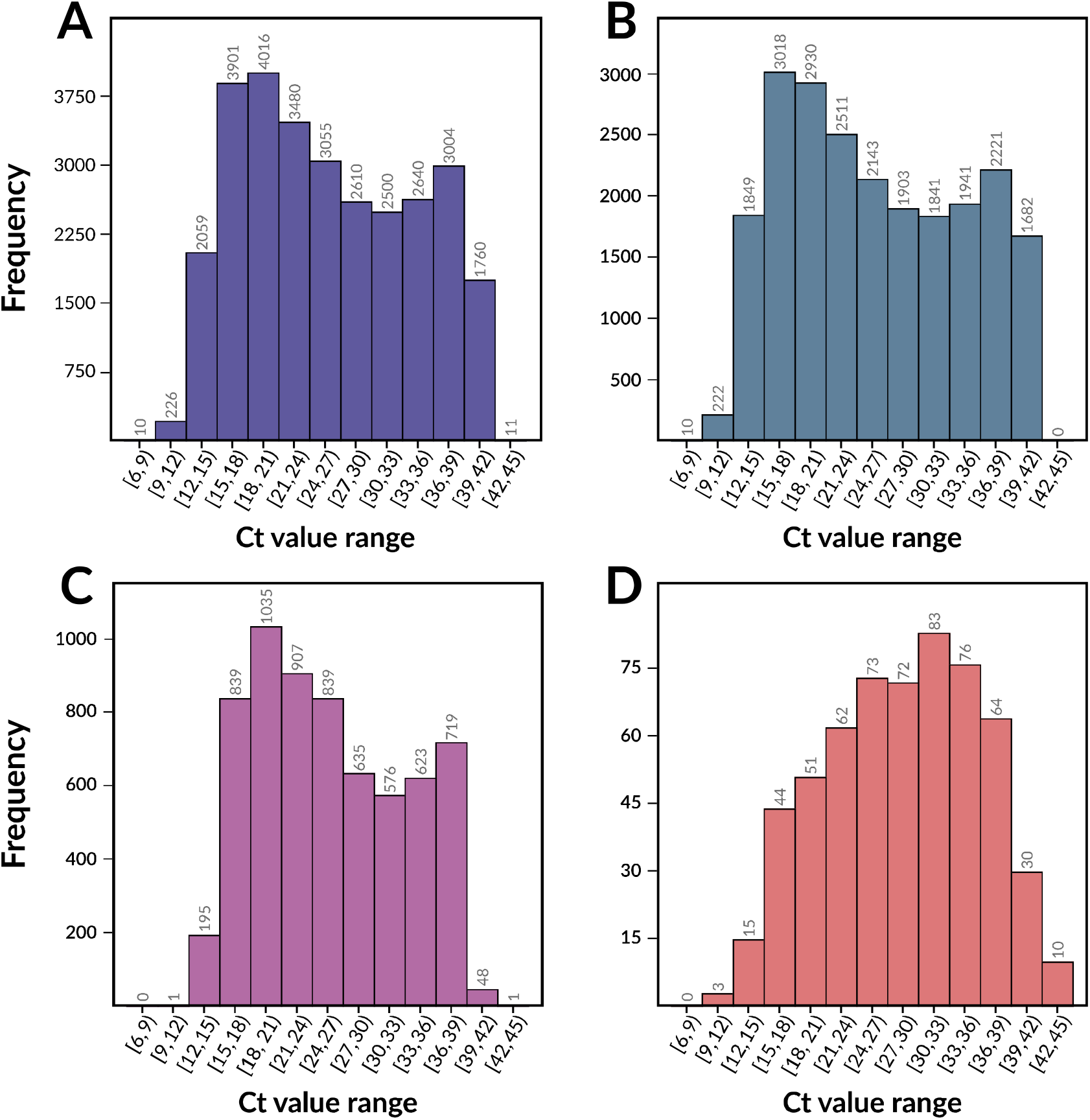
Histograms showing the distributions of Ct values in the dataset. Ct values were sorted into bins of 3 with an inclusive lower bound and exclusive upper bound. **A**: The distribution of Ct values across all detection instruments. **B**: The distribution of Ct values in the genomes from the Alinity detection instrument. **C**: The distribution of Ct values in the genomes from the Panther detection instrument. **D**: The distribution of Ct values in the genomes from the Cepheid detection instruments.

Since the objective was to determine if models built from genome sequence data could predict Ct values, we constructed feature matrices using the frequencies of all unique nucleotide 10-mers from each genome. Three types of regression models were considered to determine the best modeling approach for this study: random forest, XGBoost, and logistic regression. The logistic regression model failed to converge using max iteration values ranging from 50 to 1,500. XGBoost and random forest did not perform statistically differently using default parameters, or when comparing random forest to a set of informed parameters for XGBoost published in similar studies (43, 48) (**Table S3**). For this reason, we chose to proceed with random forest, conducting parameter sweeps to optimize the predictions (Tables S4 and S5). The accuracy of the random forest model built from all 29,272 genomes was assessed through 5-fold cross validation. The average R^2^ of the model was 0.521 ± 0.010 with an average root mean squared error (RMSE) of 5.704 ± 0.034 (Table 1), demonstrating that the model is able to detect the presence of a modest signal in the genomes.

**Table 1.** Ct value prediction results for a model built using all detection instruments and separate models for each instrument.

| Model | Genomes | R<sup>2</sup>* | RMSE* |
| --- | --- | --- | --- |
| All Instruments | 29,272 | 0.521 ± 0.010 | 5.704 ± 0.034 |
| Alinity only | 22,271 | 0.535 ± 0.014 | 5.827 ± 0.075 |
| Panther only | 6,418 | 0.417 ± 0.022 | 5.475 ± 0.113 |
| Cepheid only | 583 | 0.226 ± 0.039 | 6.787 ± 0.297 |

\*Data are reported as the average over all 5 folds with 95% confidence intervals.

### Models built from all detection instruments are more robust

In order to assess how the model performance is influenced by each detection instrument, separate models were trained using the genomes corresponding to each instrument. The model trained on the most common instrument in the dataset, Alinity with 22,271 genomes, had an R^2^ score of 0.535 ± 0.014 and an RMSE of 5.827 ± 0.075 and was similar to the model trained with all instruments (Table 1). The models trained on only Panther and only Cepheid genomes demonstrated lower performances. The Panther-only model (6,418 genomes) had an R^2^ score of 0.417 ± 0.022 and an RMSE of 5.475 ± 0.113, and the Cepheid-only model (583 genomes) had an R^2^ score of 0.226 ± 0.039 and an RMSE of 6.787 ± 0.297. The reduced accuracy is likely due to the smaller training dataset sizes with these instruments compared to the model trained on Alinity genomes.

Similarly, we evaluated how well the all-instrument model performed on the test data from each instrument (Table 2). In this case, with all of the instruments in the training set, the R^2^ and RMSEs on each instrument are more similar. As expected, the highest accuracy was observed when testing the all-instrument model on Alinity genomes, as Alinity made up the highest portion of the training set (R^2^ = 0.527 ± 0.007 and RMSE = 5.869 ± 0.036), versus Panther (R^2^ = 0.503 ± 0.024 and RMSE = 5.052 ± 0.135) and Cepheid (R^2^ = 0.366 ± 0.087 and RMSE = 6.094 ± 0.477). These data show that when all of the instruments are used together, the predictions for Panther and Cepheid samples improve.

**Table 2.** R^2^ and RMSE scores for each instrument in the test set using the model that was trained using data from all instruments.

| Instrument | Test Set Size | Fraction of Training Set | R<sup>2</sup>* | RMSE* |
| --- | --- | --- | --- | --- |
| Alinity | 4,442.2 | 0.761 | 0.527 ± 0.007 | 5.869 ± 0.036 |
| Panther | 1,292.4 | 0.219 | 0.503 ± 0.024 | 5.052 ± 0.135 |
| Cepheid | 120.4 | 0.020 | 0.366 ± 0.087 | 6.094 ± 0.477 |

\*Data are reported as the average over all 5 folds with 95% confidence intervals.

### Assessing model accuracy by Ct value

In order to understand how well the model worked over the range of Ct values, we plotted the predicted versus actual values for the all-instrument model for a single fold of the 5-fold cross validation (**Figure 3**). Two large clusters can be observed in the plot corresponding with the highest and lowest Ct values. The higher Ct values consist primarily of variants B.1 and B.1.2 and the lower Ct values consist primarily of Omicron and Delta. Although there are obvious outliers in the top left and bottom right sections of the plot, the trend is roughly diagonal and reflects the model’s predictiveness. When separate models are computed based on each instrument, the results of the Alinity-only and Panther-only models are similar to the all-instrument model, and the diagonal pattern is mostly lost in the Cepheid-only model (**Figure S1**). To assess the bounds of the accuracy of the model, we binned predicted and actual Ct values into bins of size 3 cycles, plotting the results in a confusion matrix heatmap (**Figure 4**). Like the scatterplot, the predictions form a roughly diagonal pattern, corresponding to the accurate predictions. We observe errors in several notable places including Ct values of 6-12 being predicted as 18-21, as well as incorrect predictions in Ct values >39. In the center of the plot, the fraction of correct predictions is lower, corresponding with the lower density of data points. When the predictions are generated using separate models for each instrument, the Alinity-only results are nearly identical to the all-instrument results (**Figure S2**). For the Panther-only and Cepheid-only models, we observe a propensity to over predict Ct values in the range of 21-24, and 21-27, respectively.

**Figure 3.**
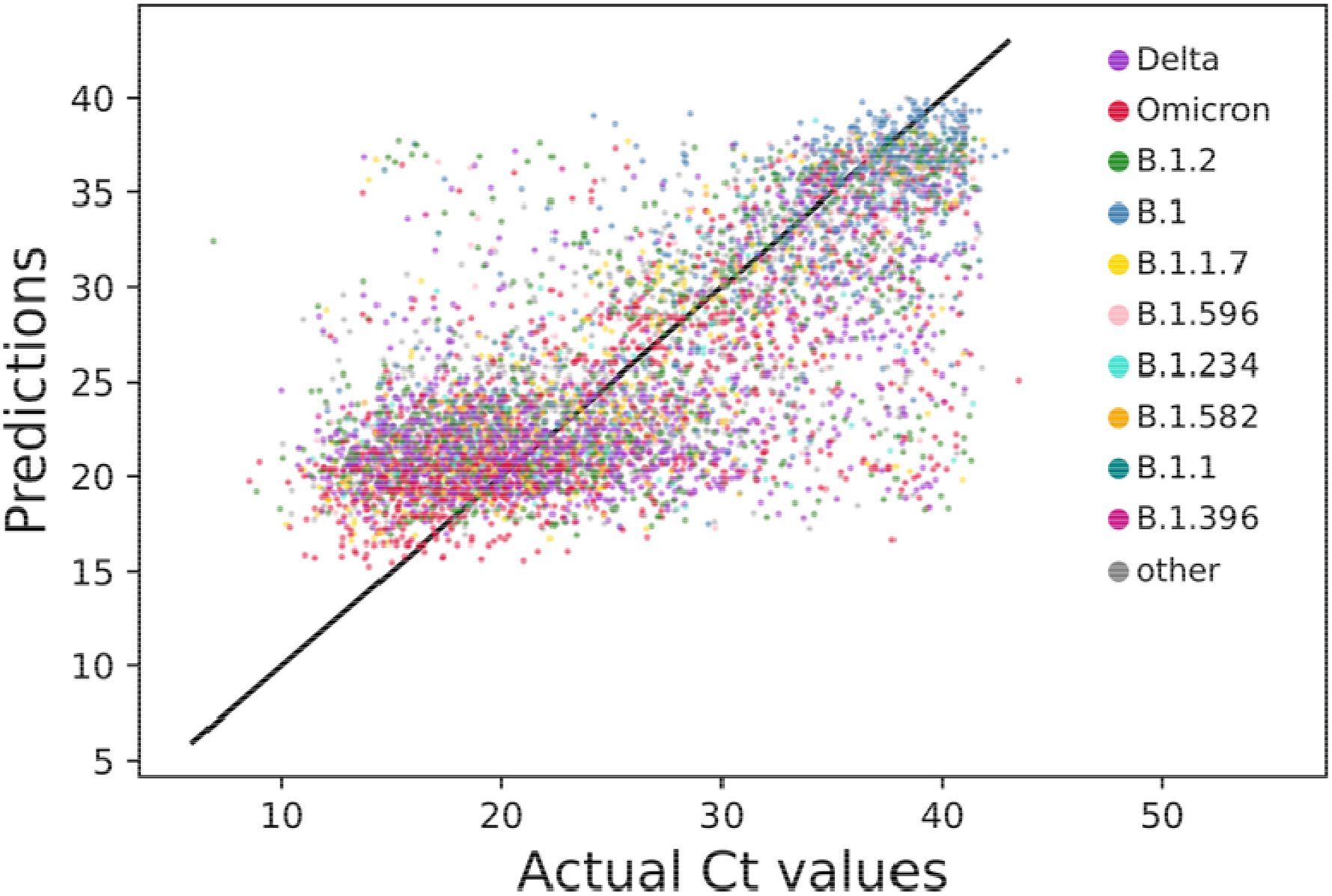
A scatterplot of predicted versus actual Ct values using the model trained on all instruments and a single fold of the 5-fold cross validation. Points are colored by variant with the 10 most frequently occurring variants colored via the key shown in the right, and samples of other variants colored gray. The line y = x is shown across the center diagonal of the figure for reference.

**Figure 4.**
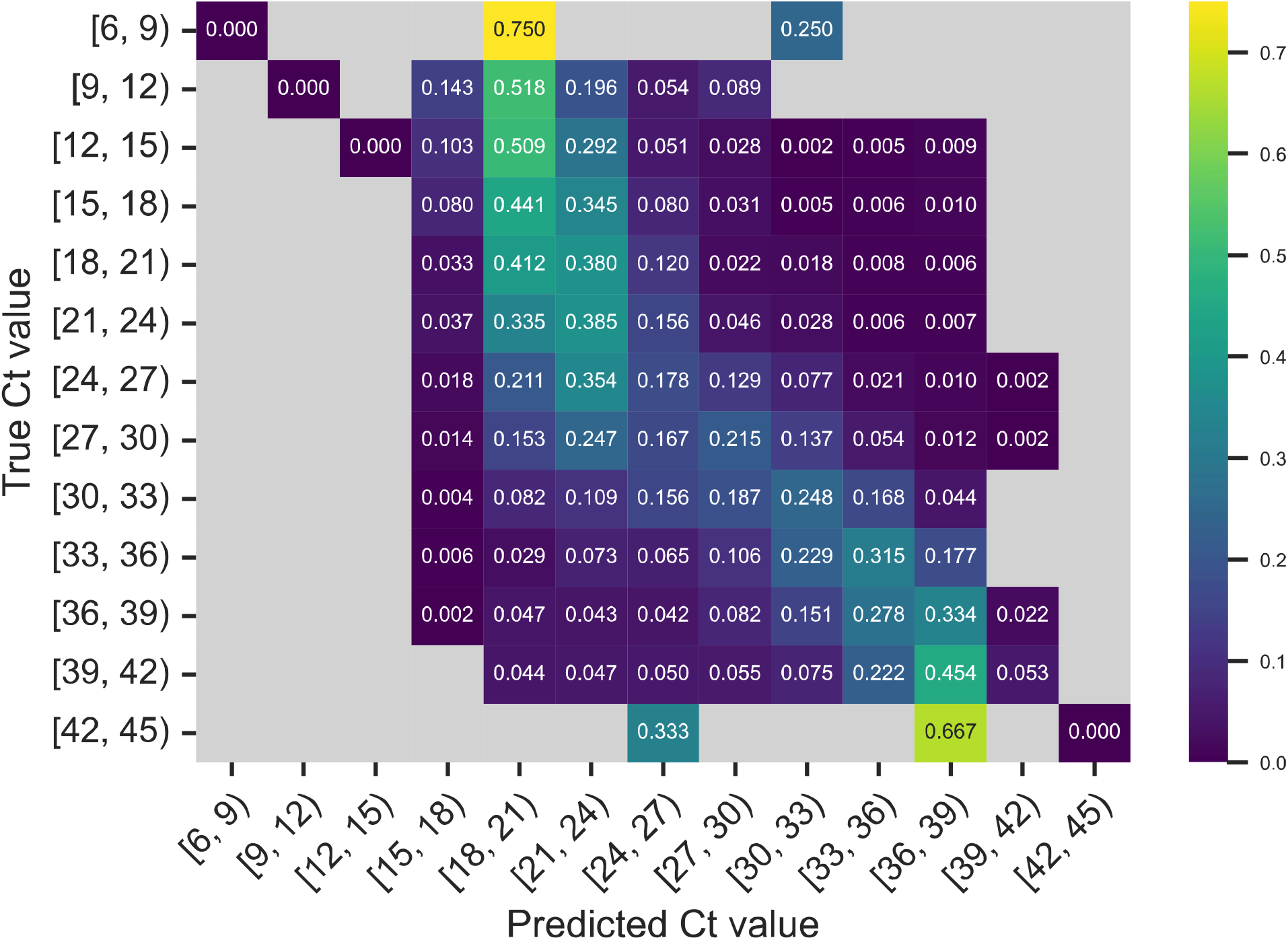
Normalized confusion matrix for a model trained on all instruments in a single fold with an 80/20 split. Model predictions were binned into Ct value ranges of size 3 with an inclusive lower bound and exclusive upper bound. Coloring and values in each cell represent the fraction of the actual Ct values predicted in the given interval. Empty cells with no predictions or predictions are gray.

Although the R^2^ value of the all-instruments regression model indicates that the model has predictive value, it is difficult to understand this in terms of the model’s overall accuracy. To approximate the overall accuracy of the model, we computed the accuracy of the model within a range of Ct values. Overall, the model exceeds 50% accuracy at a Ct value interval of approximately 3.5 cycles and exceeds 75% accuracy at an interval of 6 cycles (**Figure S3**). However, we note that this approximation of model accuracy does not account for error rates or discordance between detection instruments, which can be rather large (8, 49, 50).

As another approach, we clustered all samples with Ct values less than or equal to 24 to create a low Ct value set, and all Ct values greater than or equal to 36 to create a high Ct value set. We then built a random forest classifier to predict the high and low sets using the model parameters and cross validation described above. The resulting model had an F1 score of 0.890 ± 0.003, indicating that this coarse classification between high and low Ct values is fairly straight forward.

Because the data set almost entirely comprised of samples from outpatients (64%), inpatients (29%), and ICU patients (6%), and these sets are not balanced, we computed the R^2^ and RMSE scores for each category using the all-instrument model. Overall, the outpatient samples have a slightly lower median Ct value of 24.0 compared with 26.0 and 26.5 for the inpatient and ICU sets, respectively (**Table S8**). The model performed better over the outpatient set (R^2^ = 0.549 ± 0.013) compared with the inpatient (R^2^ = 0.453 ± 0.019) and ICU sets (R^2^ = 0.473 ± 0.030). This indicates that either the inpatient and ICU samples are a source of error due to their imbalance, genuine biological differences in these patient populations, or differences in sample preparation or handling.

### Predicting Ct values for a new VOC

Due to the importance of being able identify VOCs early, we evaluated the ability of a model to predict the Ct values of a newly emerging variant. To do this, we trained an all-instrument model but removed the Omicron genomes from the training set to simulate a model created before the emergence of Omicron. We then evaluated this model on a test set containing varying amounts of Omicron genomes. The model trained without Omicron genomes was unable to predict Ct values for Omicron genomes, likely due both to the greater number of mutations in Omicron (51) and the difference in nature of these mutations compared with previous VOCs (52) (**Table S9**). These results demonstrate that for a model designed in this way to be able to predict the Ct values of a new variant, some exposure to that variant or its mutations is needed in the training set.

In order to evaluate how much data from a new variant the model would need, we created training sets with increasingly more Omicron genomes. Models were trained on sets containing 0–15% Omicron genomes to simulate the increasing availability of genome sequences of an emerging variant. The performance of each of these models was evaluated using a test set containing the same percentage of Omicron as the respective training set. Furthermore, we split the test set by variant and evaluated the model’s accuracy on only Omicron genomes and on all non-Omicron genomes. Overall, there was no statistically significant difference in accuracy on the non-Omicron portion of the test set among models trained with different percentages of Omicron genomes (**Table S9**). The R^2^ of the Omicron-only test set increased as more Omicron genomes were included in the training set. An initially large increase in the R^2^ score for the Omicron-only test set was observed going from 0% Omicron in the training set (R^2^ = −1.069 ± 0.084) to 1% (R^2^ = 0.291 ± 0.025) (**Figure 5**). The R^2^ continued to increase more gradually going to 15% Omicron, (0.459 ± 0.030), with the RMSE gradually decreasing. Both the R^2^ and RMSE of models trained on increasing percentages of Omicron genomes demonstrated a logistic pattern with an initial large increase in accuracy that becomes more gradual. These results demonstrate that the model is able to quickly extract patterns in the genomic data to predict viral load as it is exposed to more genomes of emerging variants.

**Figure 5.**
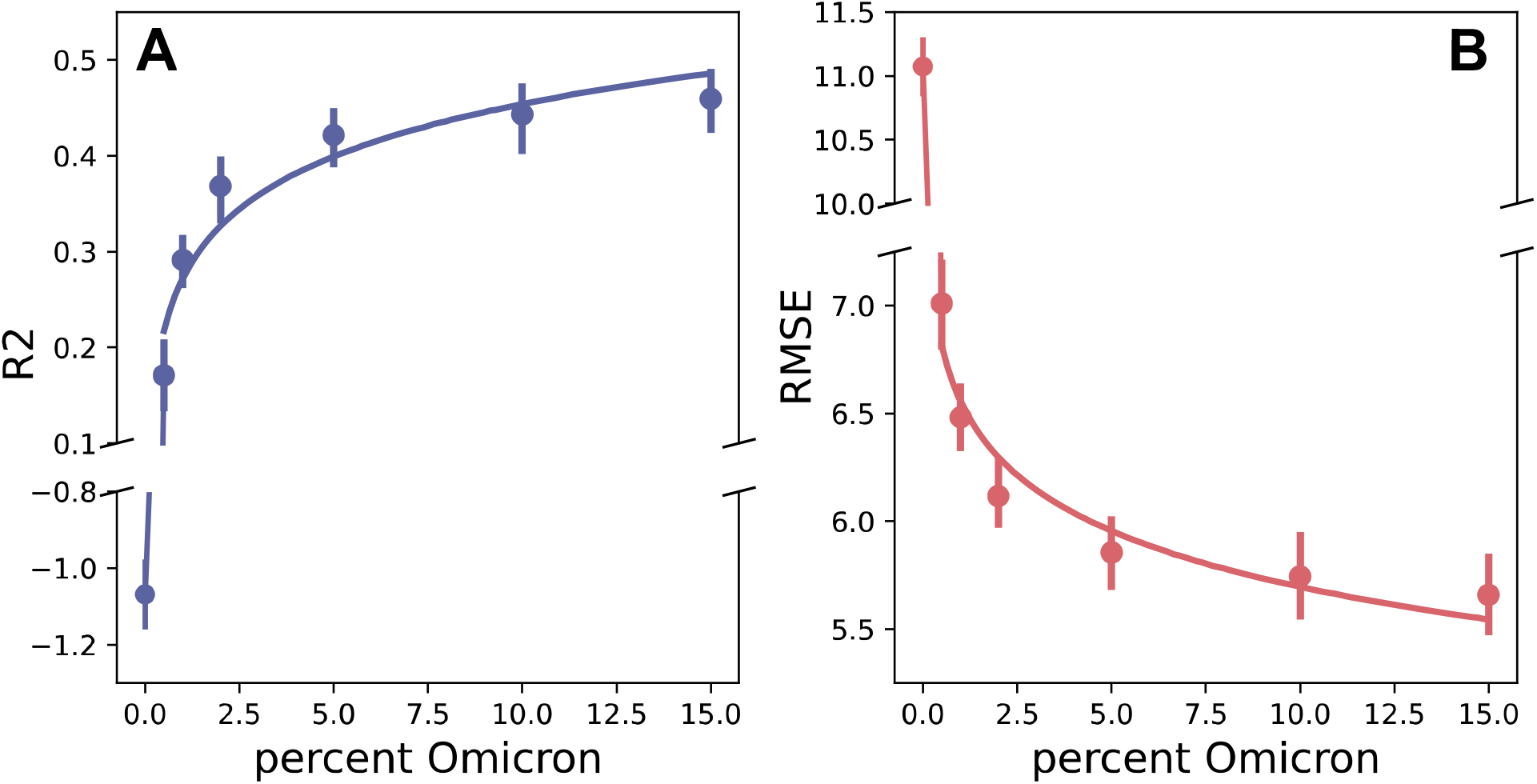
The accuracy of models trained with increasing percentages of Omicron genomes and tested on a set of only Omicron genomes. Results were calculated as the average across 5-folds with an 80% train and 20% test split with 95% confidence intervals. A logistic regression curve was plotted for Omicron percentages 0.5 to 15 with a 95% confidence interval. **A**: the R^2^ scores of the models. **B**: the RMSEs of the models.

### Feature Importance

In order to gain insight into the genomic regions that the model is using to predict Ct values, we chose to examine the important k-mers that were chosen by the all-instrument model that was trained on all variants. Since the feature importance function for random forest reports Gini impurity, it is possible to have decision nodes (k-mers) with very low impurity that are rarely used by the model. In essence, these are k-mers that are rare but split the data set very well. Indeed, when we examined the data in this way, this was our observation.

As a different approach, we chose to assess the decision nodes that are visited most frequently in the random forest, reasoning that these may contain information about the relationship between the k-mers and the predicted Ct values. To do this, we computed all decision paths for each genome in the training set. We then filtered these for the nodes that were encountered most frequently across all of the genomes and trees, and for which there was a significant difference in the average Ct values of the genomes with and without the k-mer. The k-mers that occurred most frequently relative to the background were then plotted based on their corresponding positions in the Wuhan-Hu-1 reference genome (**Figure 6**). Overall, we observe that the model is using many k-mers from across the entire genome to make the predictions. However, like Gini impurity, a node that is frequently encountered in the decision path does not need to occur in many genomes because the decision trees, which make up a random forest, are greedy (53).

**Figure 6.**
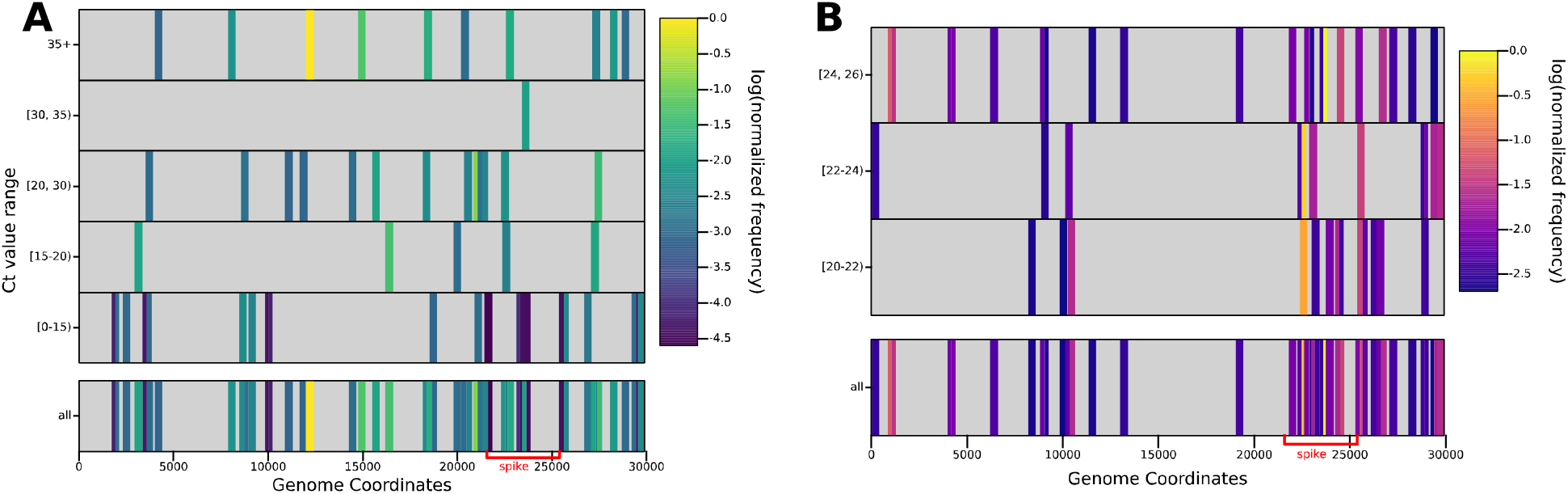
Heatmaps depicting relevant k-mers used by the all-instrument model. The horizontal axis depicts the location of each k-mer on the SARS-CoV-2 genome and the vertical axis depicts the average Ct value range for the genomes containing the k-mer, with the bottom row showing all features combined. Each bar represents a k-mer (not drawn to scale) and the coloring representing the number of times that the k-mer is used in the in the decision paths for the training set genomes (log normalized). **A**: the most frequently occurring features in all decision paths, where there is a statistically significant difference between the average Ct values of the genomes with and without the k-mer, that are used above a baseline threshold. **B**: The most frequently occurring features in all decision paths, where there is a statistically significant difference between the average Ct values for genomes with and without the k-mer, and the k-mers occur in at least 10% of the genomes.

To refine the search, we filtered the set of significant k-mers for those occurring in at least 10% of the genomes. When we did this, we observed a clustering of 15 significant k-mers in the spike protein (**Figure 6**, **Table S10**). These k-mers correspond with important positions that are hallmarks of various VOCs, six of which occur in the receptor binding domain. Several notable regions in the spike protein that are spanned by these k-mers include: Y144- and Y145-, G339D, K417N/T, L452R/Q, N501Y, and P618H/R, which are hallmarks of various VOCs including Alpha, Delta, and Omicron. In these instances, the model has chosen k-mers in regions that are important in the evolution of the VOCs.

However, it is clear that the model cannot simply rely on the major VOC SNPs to predict the Ct values, and that it needs the more subtle non-linear relationships between many features in order to achieve the R^2^ score that we observe.

## Discussion

Having the ability to identify and predict clinical characteristics of SARS-CoV-2 is of paramount importance for epidemiology and infection control. Although they are rarely used in modeling studies, Ct values tend to correlate with viral loads and offer insight into the contagiousness and transmissibility of the virus (9–11). In this study, we used genome sequence data to predict Ct values for a collection of 29,272 SARS-CoV-2 clinical samples. After evaluating a variety of algorithms and conducting parameter sweeps, our best model was a random forest regressor that could predict Ct values with an R^2^ of 0.521 and an RMSE of 5.7. This indicates the presence of a modest signal in the genome sequence data that the models are identifying and using to make the predictions.

In this study, we chose a machine learning approach in order to predict Ct values. The machine learning methods that were used in this study all enable a rather seamless examination of the top features (k-mers), which we deemed to be important for understanding how the model worked. Likewise, we chose a longer k-mer length so that we could easily identify the genomic coordinates of each k-mer. When we evaluated the top features that occurred in the most genomes, we observed a cluster of k-mers occurring in the spike protein. Many of these k-mers span regions of the spike protein that are hallmarks of previous VOCs, supporting the notion that many of these mutations have an impact on viral load (26). However, it is clear from our analysis of the Gini impurity and frequently encountered nodes in the decision paths of the forest that many k-mers are required to obtain the R^2^ and RMSE that we observe for the all-instrument model. This is unsurprising for two reasons. First, the random forest algorithm is greedy and will choose the k-mers that differentiate the high and low Ct values, even if they occur rarely in the data set. Second, since this is a regression problem aimed at predicting Ct values along a continuum, more k-mers are required to suss out the subtle step-wise differences between samples.

Predicting Ct values turned out to be a challenging modeling problem because of the nature of the clinical data set. The clinical data were imbalanced in at least two ways. First, the use of the detection instruments was not uniform, with the Alinity instrument comprising 76% of the samples in the collection followed by Panther (22%) and Cepheid (2%). Secondly, the patient cohort was comprised of approximately 64% outpatients, 29% inpatients, and 6% ICU patients, in various stages of disease progression. In both cases, it is clear that the less represented sets create a source of error model, with these subsets having lower R^2^ values and higher RMSEs. It is possible that sampling differences may account for some of the error. For instance, the Cepheid instruments are more frequently used for critical patients because they have a faster turnaround time of 1 hour compared with 4-6 hours for the other instruments. Ultimately, we showed that given adequate data, separate models could be built for these groups, and may be the most appropriate solution to this problem.

Likewise, we found that it was important to have high quality genome sequence data. Over the course of the pandemic, there have been fluctuations in overall sequence quality due to the use of various sequencing methods and primer kits (54). To overcome this, we chose to filter all genomes for those having less than 500 ambiguous nucleotide characters, and we also needed to mask the ends and 56 positions within the spike protein-encoding gene in order to prevent the models from using variation that was due to poor sequence quality rather than true variation. As approaches with higher accuracy and fewer errors are used, we anticipate that the signal will become stronger.

Assessing the overall accuracy of the model was challenging because the highest achievable accuracy is limited by the error rate of the detection instruments. Several studies have published comparisons of SARS-CoV-2 clinical detection instruments, reporting good agreement across instruments, as well as strong correlation in Ct or cycle number (CN) values (49, 50, 55). However, there is an error rate when performing technical replicates from the same sample on the same device, and from comparing the results from the same sample across different devices. For example, Perchetti and colleagues performed a comparison of the Ct values from the Alinity *m* and Panther Fusion instruments for the purpose of validating the Alinity *m* (49). Using 30 matched positive samples, they reported an average Ct difference of approximately 0.7 between two replicates of the Alinity *m* and an average Ct value difference of approximately 2 between matched samples from Alinity *m* and Panther Fusion. Although the number of samples compared in that study was small, we can use their results to roughly establish the expectation that a perfect model for predicting Ct values would have a margin of error in the range of at least 0.7-2.0 cycles. However, this is probably a low estimate considering that larger differences have been reported for other instruments(8, 49, 50), and accounting for the large number of samples in our study with Ct values above 30, which are likely to have greater detection error. For this reason, the accuracy of the model presented in this study is likely understated.

Having the ability to identify new variants early, before there is community transmission, is necessary for controlling the SARS-CoV-2 pandemic. We attempted to simulate this by training models on prior variants and testing them on Omicron genomes. Using this approach, we found that approximately 5% of the genomes in the training set needed to be from Omicron in order to accurately predict Ct values for Omicron genomes, i.e., the mutations of Omicron needed to be learned. This is somewhat unsurprising considering the large difference in the mutational profiles of Omicron versus previous variants (51, 52). However, the small percentage of Omicron genomes that is required to begin to achieve accurate models is encouraging and provides a baseline for improvement. Future studies aimed at more accurately predicting Ct values may be improved by incorporating positional information from alignments, epitopes, and other experimental data as features. For instance, a recent study by Mushegian and colleagues noted a relationship between minor variants and Ct values, with higher Ct value samples having more minor sequence variants (56).

In summary, this study demonstrates it is possible to use SARS-CoV-2 genome sequence data to predict Ct values. This provides a modeling framework and baseline for improving the identification of VOCs and the prediction of clinical characteristics of SARS-CoV-2 variants, which are important for future infection control efforts.

## Data Availability

All data produced in the present work are contained in the manuscript

## Abbreviations

BV-BRC: Bacterial and Viral Bioinformatics Resource Center
Ct: cycle threshold
VOC: variant of concern
RMSE: root mean square error
RT-PCR: reverse transcription-polymerase chain reaction
XGBoost: extreme gradient boosting

## Acknowledgements

We thank Robert Wisniewski and the BV-BRC and Houston Methodist teams for their helpful input. We thank Alma Amaya, Akanksha Batajoo, Jessica Cambric, Ryan Gadd, Nicole Kanellopoulos, Shelby Kvinta, Regan Mangham, Eleanor Nichols, Jordan Pachuca, Sindy Pena, Kristina Reppond, Matthew Ojeda Saavedra, Madison Shyer, and Rashi Thakur, and Bob Olson for technical assistance. LD was funded by the Northwestern-Argonne Institute of Science and Engineering (NAISE) Summer Research Experience Program supported by Northwestern University’s Office for Research. JJD and MN were supported by the National Institute of Allergy and Infectious Diseases, National Institutes of Health, Department of Health and Human Services [75N93019C00076 to PI Rick Stevens]. This work was also supported by Discovery Partners Institute award [PRJ1009544] to JD. PC, SWL, RJO, and JMM were supported by the Houston Methodist Academic Institute Infectious Diseases Fund.

## Conflict of Interest

The authors of this work declare no conflicts of interest.

## Notes

### Competing Interest Statement

The authors have declared no competing interest.

### Author Declarations

All samples were collected with patient consent and Institutional Review Board approval from the Houston Methodist Research Institute (IRB1010-0199).

