## Supplemental Figures for "Using Genome Sequence Data to Predict SARS-CoV-2 Detection Cycle Threshold Values"

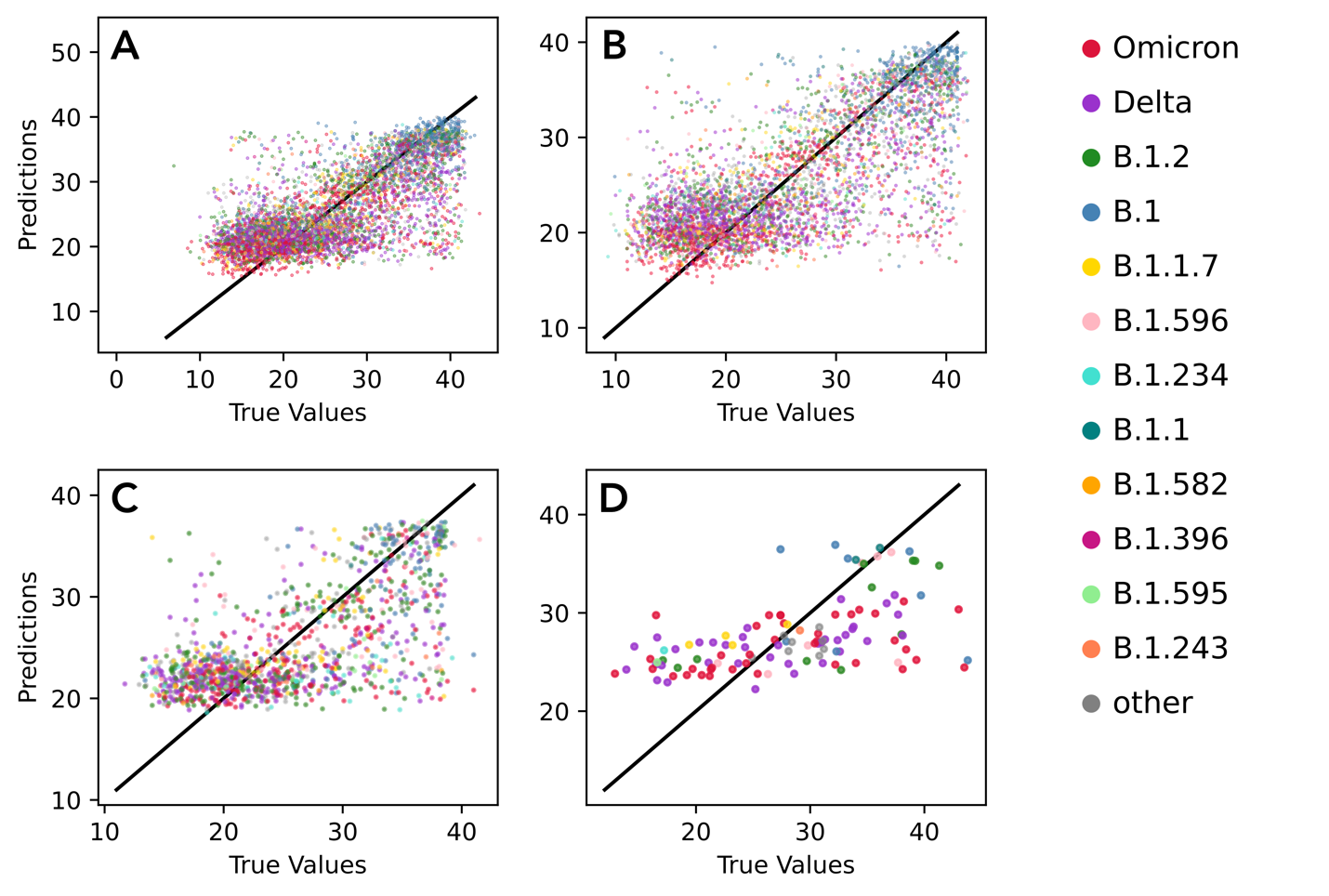

**Figure S1.** Scatterplots of predicted versus actual Ct values. Scatterplots were constructed for models trained and tested on the following instruments: **A:** All instruments. **B:** Alinity, **C:** Panther, **D:** Cepheid. Points are colored by variant with samples of the 10 most frequently occurring variants colored via the key shown in the right, and samples of other variants colored gray. The line y = x is shown across the center diagonal of the figure for reference. Data are from a single fold of a 5-fold cross validation for each model.

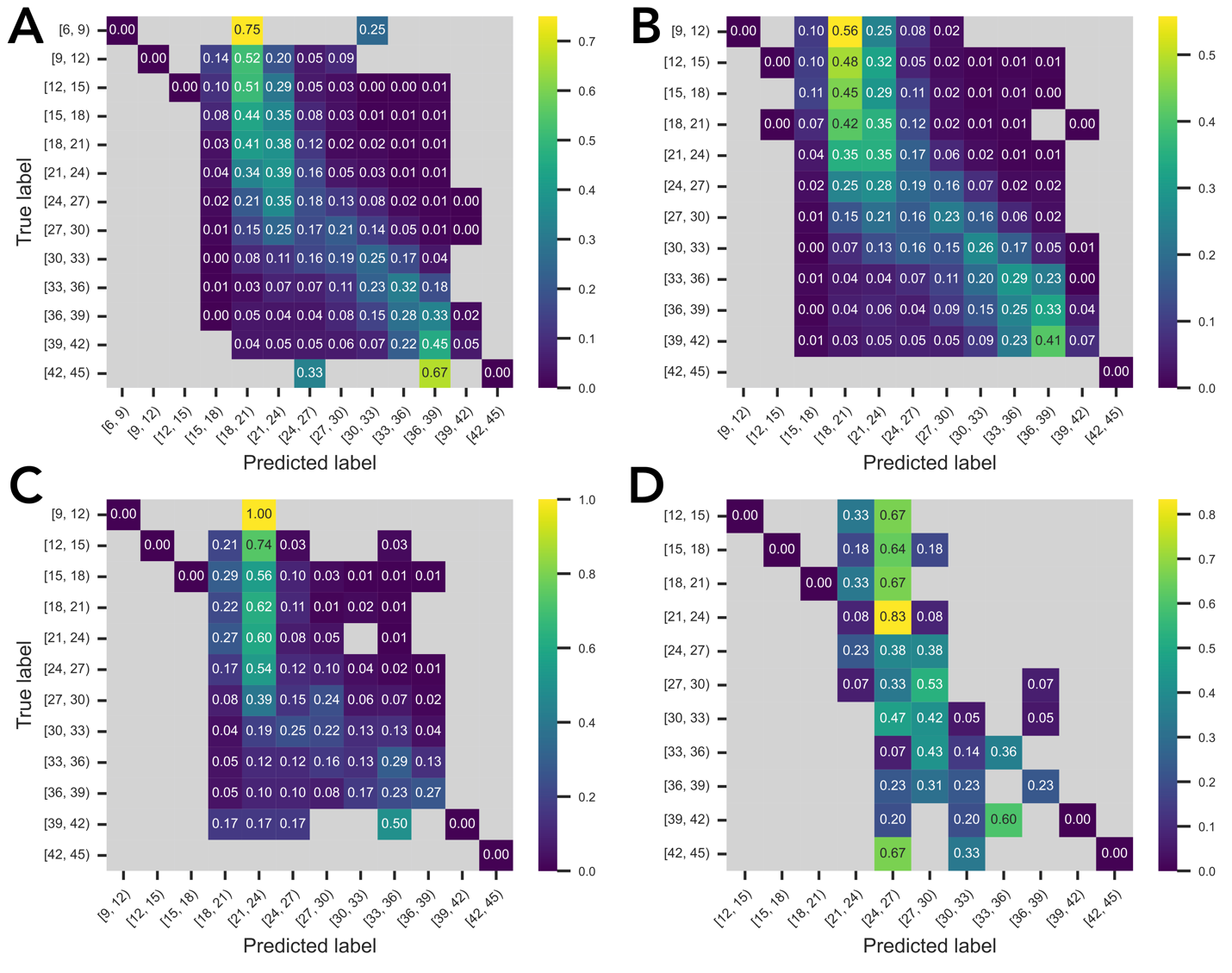

**Figure S2.** Confusion matrix for models using a single fold of the 5-fold cross validation. Predictions were binned into Ct value ranges of 3 cycles with an inclusive lower bound and exclusive upper bound. Coloring and values in each cell represent the fraction of the actual Ct values predicted in the given interval. Empty cells with no predictions or actual values in that range are gray. Confusion matrices were constructed for models trained and tested on the following instruments: **A:** All instruments. **B:** Alinity-only, **C:** Panther-only, **D:** Cepheid-only.

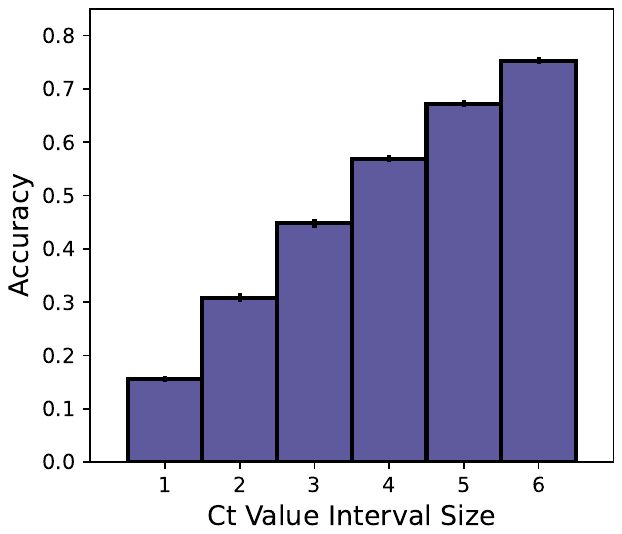

**Figure S3.** The accuracy of the all-instrument model within a series of Ct value intervals. The accuracy for a given interval *n* was calculated as the fraction of predictions that were within *n* or less cycles from the actual value. All results were calculated as the average across a 5-fold cross validation with an 80% train 20% test split. Error bars depict the 95% confidence intervals.
